# Predicting the epidemic trend of COVID-19 in China and across the world using the machine learning approach

**DOI:** 10.1101/2020.03.18.20038117

**Authors:** Mengyuan Li, Zhilan Zhang, Shanmei Jiang, Qian Liu, Canping Chen, Yue Zhang, Xiaosheng Wang

## Abstract

**Background:** Although COVID-19 has been well controlled in China, it is rapidly spreading outside the country and may have catastrophic results globally without implementation of necessary mitigation measures. Because the COVID-19 outbreak has made comprehensive and profound impacts on the world, an accurate prediction of its epidemic trend is significant. Although many studies have predicted the COVID-19 epidemic trend, most have used early-stage data and focused on Chinese cases.

**Methods:** We first built models to predict daily numbers of cumulative confirmed cases (CCCs), new cases (NCs), and death cases (DCs) of COVID-19 in China based on data from January 20, 2020, to March 1, 2020. Based on these models, we built models to predict the epidemic trend across the world (outside China). We also built models to predict the epidemic trend in Italy, Spain, Germany, France, UK, and USA where COVID-19 is rapidly spreading.

**Results:** The COVID-19 outbreak will have peaked on February 22, 2020, in China and will peak on May 22, 2020, across the world. It will be basically under control in early April 2020 in China and late August 2020 across the world. The total number of COVID-19 cases will reach around 89,000 in China and 6,126,000 across the world during the epidemic. Around 4,000 and 290,000 people will die of COVID-19 in China and across the world, respectively. The COVID-19 outbreak will have peaked recently in Italy and will peak in Spain, Germany, France, UK, and USA within two weeks.

**Conclusion:** The COVID-19 outbreak is controllable in the foreseeable future if comprehensive and stringent control measures are taken.

## Background

Although the spread of COVID-19 caused by the 2019 novel coronavirus (SARS-CoV-2) is slowing down in China, it is rapidly growing outside the country, and how it will evolve remains unclear. So far, SARS-CoV-2 has infected more than 2,000,000 people and led to nearly 135,000 deaths worldwide. The COVID-19 outbreak has been declared a pandemic and is expected to cause one of the most serious global public health problems in recent years.^1^ Of note, the impacts of the COVID-19 epidemic are not only limited to global public health but also affect the global economy, geopolitics, culture, and society. Thus, an accurate prediction of the epidemic trend of COVID-19 may provide valuable advice on how to effectively control its spread and relieve its major social and economic impacts. Although a number of studies have estimated the epidemic trend of the COVID-19 outbreak, most have used early-stage data and focused on cases in China.^2-8^ For example, using the SEIR (susceptible, exposed, infectious, and removed) model, Wang et al. estimated the numbers of COVID-19 cases in Wuhan, China, under the conditions of insufficient and sufficient control measures being taken.^3^ Wu et al. predicted the domestic and global spread of SARS-CoV-2 based on travel volume data during Dec 2019 and Jan 2020.^4^ Chen et al. evaluated the transmissibility of SARS-CoV-2 using the reservoir-people transmission network model.^7^ Yang et al. predicted the epidemic peaks and scales of COVID-19 using the SEIR model and machine learning to show the indispensability of the nationwide interventions starting from January 23, 2020.^8^ With COVID-19 remaining active in China and continuing to spread around the world, predicting its epidemic trend in China and across the world based on updated data is definitely needed.

In this study, based on the public data of COVID-19 cases, we predicted its epidemic trend in China and across the world. We first built a prediction model using the publicly available data for COVID cases. These data include daily numbers of cumulative confirmed cases (CCCs), new cases (NCs), and death cases (DCs) of COVID-19 in China since January 20, 2020. We trained our model using data from January 20, 2020, to March 1, 2020, and predicted the daily numbers of CCCs, NCs, and DCs. Furthermore, we predicted the epidemic trend of COVID-19 across the world (outside China) using models derived from those built based on Chinese cases.

## Methods

### Data preparation

We downloaded the statistics of confirmed COVID-19 cases in China from the National Health Committee of China (http://www.nhc.gov.cn/), including the daily numbers of CCCs, NCs, and DCs since January 20, 2020. The data for the statistics of confirmed COVID-19 cases outside China were downloaded from Worldometer (https://www.worldometers.info/coronavirus/#countries).

### Predictive models

We built models to predict daily numbers of CCCs, NCs, and DCs of COVID-19 in China based on data from January 20, 2020, to March 1, 2020, using Eureqa (Trial Version 1.24.0, https://www.nutonian.com/products/eureqa-desktop), a machine learning algorithm that can automatically build predictive models from data.^9^ We input daily numbers of CCCs and DCs and their corresponding days to obtain a formula that perfectly shaped the relationships between the variables. Due to insufficient sample sizes of the datasets for COVID-19 cases across the world (outside China) because of the recentness of the outbreak, we did not directly use Eureqa to build predictive models for the COVID-19 epidemic trend across the world. We instead derived them from the models for Chinese cases on the assumption that the COVID-19 epidemic trend outside China was similar to that in China with a time lag. Furthermore, we predicted the epidemic trend in the European and North American countries where COVID-19 is rapidly spreading, including Italy, Spain, Germany, France, UK, and USA.

## Results

### Prediction of numbers of CCCs, NCs, and DCs of COVID-19 in China

We used Eureqa to train the model to predict the daily number of CCCs of COVID-19 in China using data from January 20, 2020 to March 1, 2020. We obtained the predictive model *Y*=42641.14+30962.75×*atan*(0.16× *X* − 3.43), where *Y* is the number of CCCs and *X* is the time (days). The goodness-of-fit (*R*^2^) of the model was 0.997 on the training set. The model predicted that the number of CCCs would peak on February 22, 2020, with 76,983 CCCs, and that the epidemic curve would flatten starting from March 25, 2020, in China (Fig. 1A). This is consistent with several recent reports.^8^ We predicted that the COVID-19 epidemic would end around mid-May in China. The number of COVID-19 cases is expected to total around 89,000 when the epidemic ends in China.

**Figure 1.**
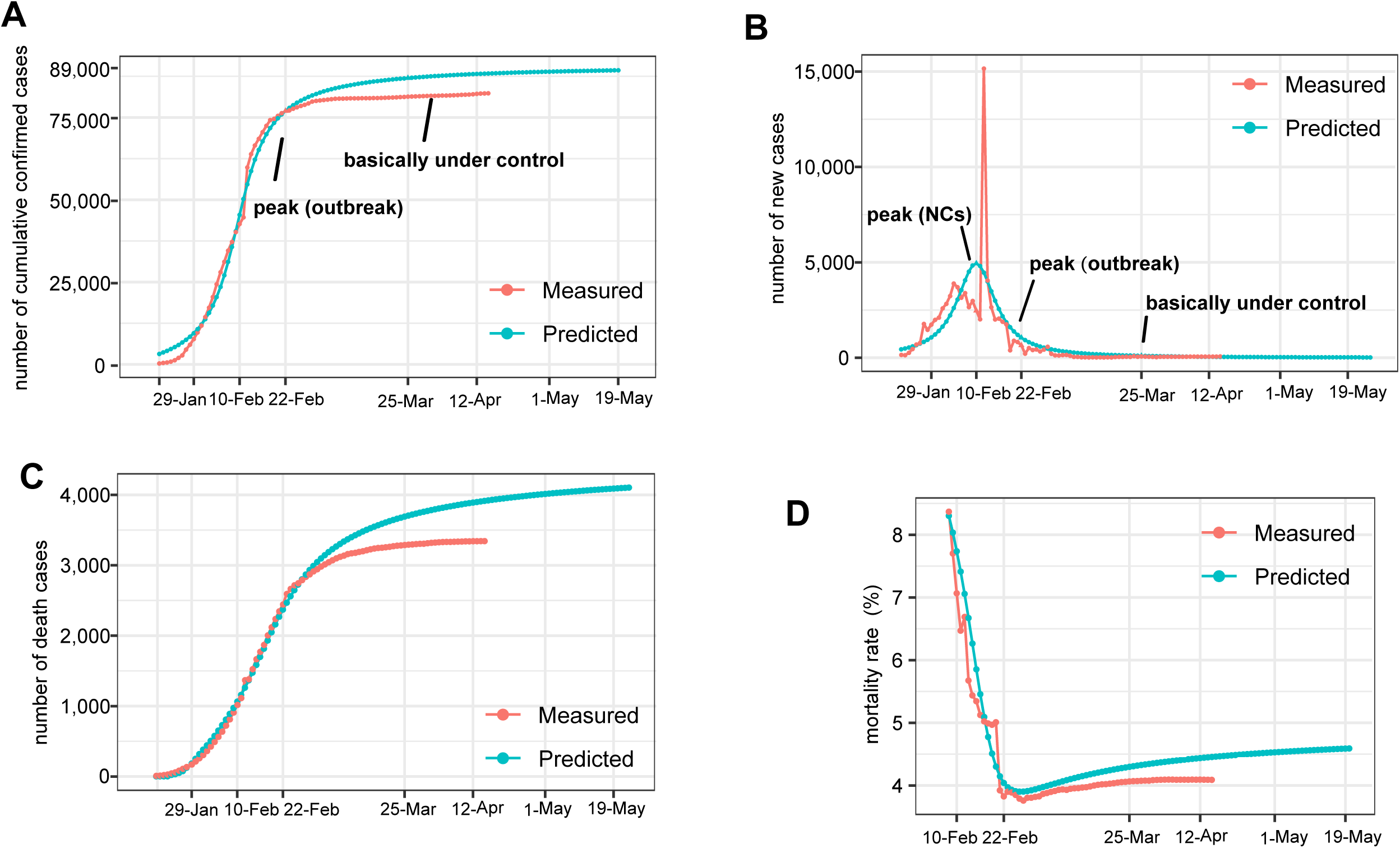
Prediction of the COVID-19 epidemic trend in China. Predicted daily numbers of cumulative confirmed cases (A), new cases (B), and death cases (C) of COVID-19 in China. D. Predicted daily COVID-19 mortality rates in China. The mortality rate on a given day is the number of death cases that day divided by the number of cumulative confirmed cases 10 days before.

On the basis of the predicted daily number of CCCs, we estimated the daily number of NCs on a given day as the number of CCCs that day minus the number of CCCs on the previous day (Fig. 1B). The number of NCs was predicted to peak at 4,944 on February 10, 2020. At the predicted peak of the COVID-19 outbreak on February 22, 2020, the number of NCs was estimated to be 1,050. After February 22, 2020, the daily numbers of NCs were expected to be less than 1,000. Starting from March 25, 2020, the daily numbers of NCs would drop to less than 100, indicating that the COVID-19 outbreak was basically under control in China.

In addition, we used Eureqa to build a model to predict the daily number of DCs in China:

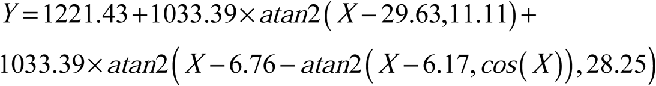

where *Y* is the number of NCs and *X* is the time (days). The model *R*^2^ was 0.998 on the training set. The model predicted around 4,000 deaths caused by COVID-19 during the epidemic in China (Fig. 1C). Furthermore, we estimated that the mortality rate for COVID-19 would be 4.2%, which is slightly higher than the mortality rate of 4.1% calculated based on the latest data on April 16, 2020. However, because the number of NCs was growing small, while the number of critical cases remained at more than 95 on April 16, 2020, the final actual mortality rate for COVID-19 in China will approach to our estimate. Furthermore, we estimated the daily COVID-19 mortality rate in China based on the daily predicted CCCs and DCs. Because the median time from the onset to critical condition is around 10 days^10^, we calculated the mortality rate on a given day by dividing the number of DCs on that day by the number of CCCs 10 days before. The daily mortality rate increases over time and becomes steady from early April 2020 (Fig. 1D).

### Prediction of epidemic trend of COVID-19 across the world

The COVID-19 outbreak across the world (outside China) has lagged behind that in China. We tried to predict its epidemic trend and found that the daily numbers of CCCs within the 14 consecutive days starting from February 27, 2020, across the world followed the same distribution as within the 14 consecutive days starting from January 26, 2020, in China (Kolmogorov− Smirnov (K−S) test, *p* = 1; Fig. 2A). Thus, we supposed that the COVID-19 epidemic across the world had a similar trend to that in China with a 32-day lag. Accordingly, based on the model to predict the daily number of CCCs of COVID-19 in China, we generated a model to predict the daily number of CCCs of COVID-19 across the world:

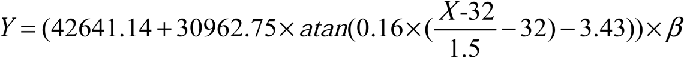

where *Y* is the number of CCCs, *X* is the time (days), and *β* (= 67.8) is the tuning parameter. The *β* value is the product of the COVID-19 control strength in China relative to the world outside China (= 1.5), the ratio of the population outside China to that in China (= 4.52), and the ratio of actual number of COVID-19 cases to number of COVID-19 cases identified (= 10).^11^ Based on this model, we predicted that the number of NCs would peak on around April 9, 2020, with around 223,000 NCs across the world (Fig. 2B). On May 22, 2020, the COVID-19 outbreak will peak across the world with the number of CCCs and NCs being around 5,746,000 and 10,000, respectively (Fig. 2B). After August 27, 2020, the daily number of NCs will be less than 1,000, indicating that the COVID-19 outbreak is basically under control across the world. The total number of COVID-19 cases is estimated to reach around 6,049,000 during the epidemic across the world.

**Figure 2.**
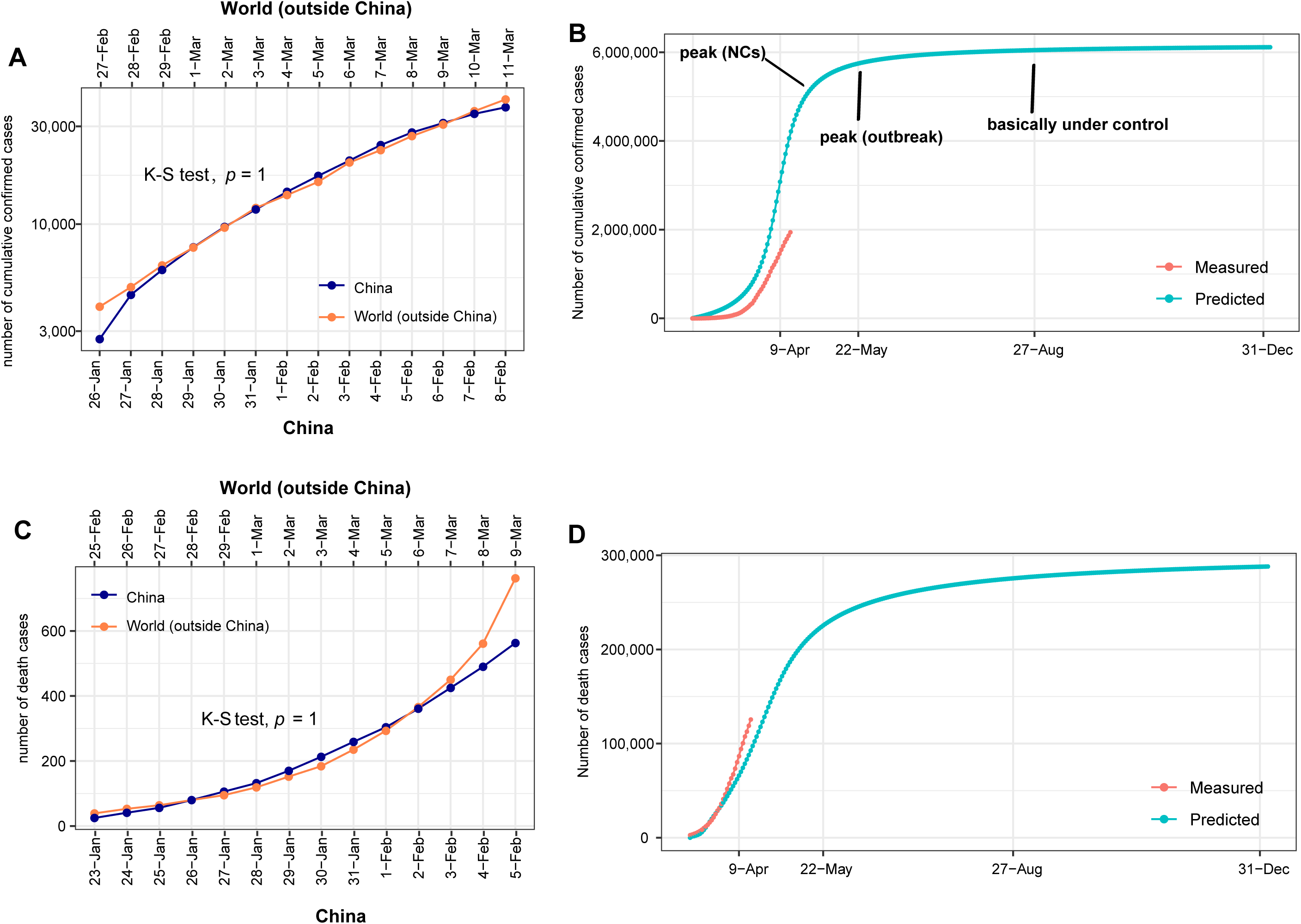
Prediction of the COVID-19 epidemic trend in the world. A. The daily numbers of cumulative confirmed cases of COVID-19 in the 14 consecutive days starting from February 27, 2020, outside China follow the same distribution as those within the 14 consecutive days starting from January 26, 2020, in China (*K-S test, p* = 1). B. Predicted daily numbers of cumulative confirmed cases and new cases of COVID-19 in the world. C. Daily numbers of death cases of COVID-19 in the 14 consecutive days starting from February 25, 2020, outside China followed the same distribution as those in the seven consecutive days starting from January 23, 2020, in China (*K-S test, p* = 1). D. Predicted daily numbers of death cases of COVID-19 outside China.

In addition, we found that the daily number of DCs in the 14 consecutive days starting from February 25, 2020, outside China followed the same distribution as those within the 14 consecutive days starting from January 23, 2020, in China (K-S test, *p* = 1; Fig. 2C). Accordingly, based on the model to predict the daily number of DCs in China, we built a model to predict the daily number of DCs across the world:

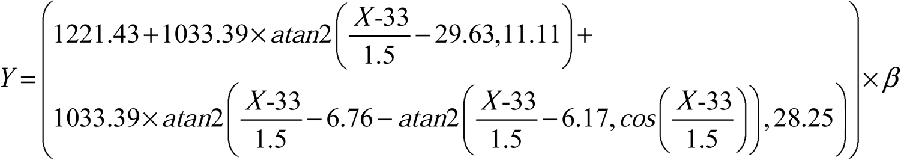

where *Y* is the number of DCs, *X* is the time (days), and *β* (= 67.8) the tuning parameter. Using this model, we predicted that a total of 276,000 people will have died of the COVID-19 disease when the outbreak is basically under control across the world (Fig. 2D). The mortality rate for COVID-19, estimated by our predictive models, will be 4.6% across the world during the COVID-19 outbreak. This number is slightly higher than that (4.2%) for China.

### Prediction of epidemic trend of COVID-19 in individual countries outside China

We observed that the daily number of COVID-19 NCs in Italy had reached its peak. Thus, we used Eureqa to train the model to predict the daily number of NCs in Italy using data from February 22, 2020 to March 26, 2020. We obtained the predictive model

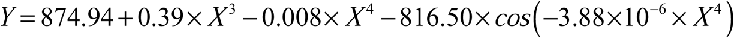

where *Y* is the number of NCs, *X* is the time (days). The model *R*^2^ was 0.9998 on the training set. Based on this model, we predicted that the number of NCs would have two peaks. The first peak would be on around March 23, 2020, with around 58,000 NCs, and the second peak would be on around March 31, 2020, with around 59,000 NCs (Fig. 3A). After April 10, 2020, the COVID-19 outbreak will be gradually brought under control in Italy. Based on this model, the total number of COVID-19 cases was estimated to be 130,000 in Italy during the epidemic. However, considering the continuous increase in testing capacity for SARS-CoV-2 infection later in Italy, we expected that the final number of COVID-19 cases would be twice this number, that is, 260,000. Furthermore, we used Eureqa to build a model to predict the daily number of DCs in Italy using data from March 5, 2020 to March 26, 2020. We obtained the predictive model *Y* = 25.33 + 2.81× *X*^2^ − 0.057 × *X*^3^, where *Y* is the number of new DCs, *X* is the time (days). The model *R*^2^ was 0.93 on the training set. It predicted around 30,000 deaths caused by COVID-19 during the epidemic in Italy (Fig. 3A).

**Figure 3.**
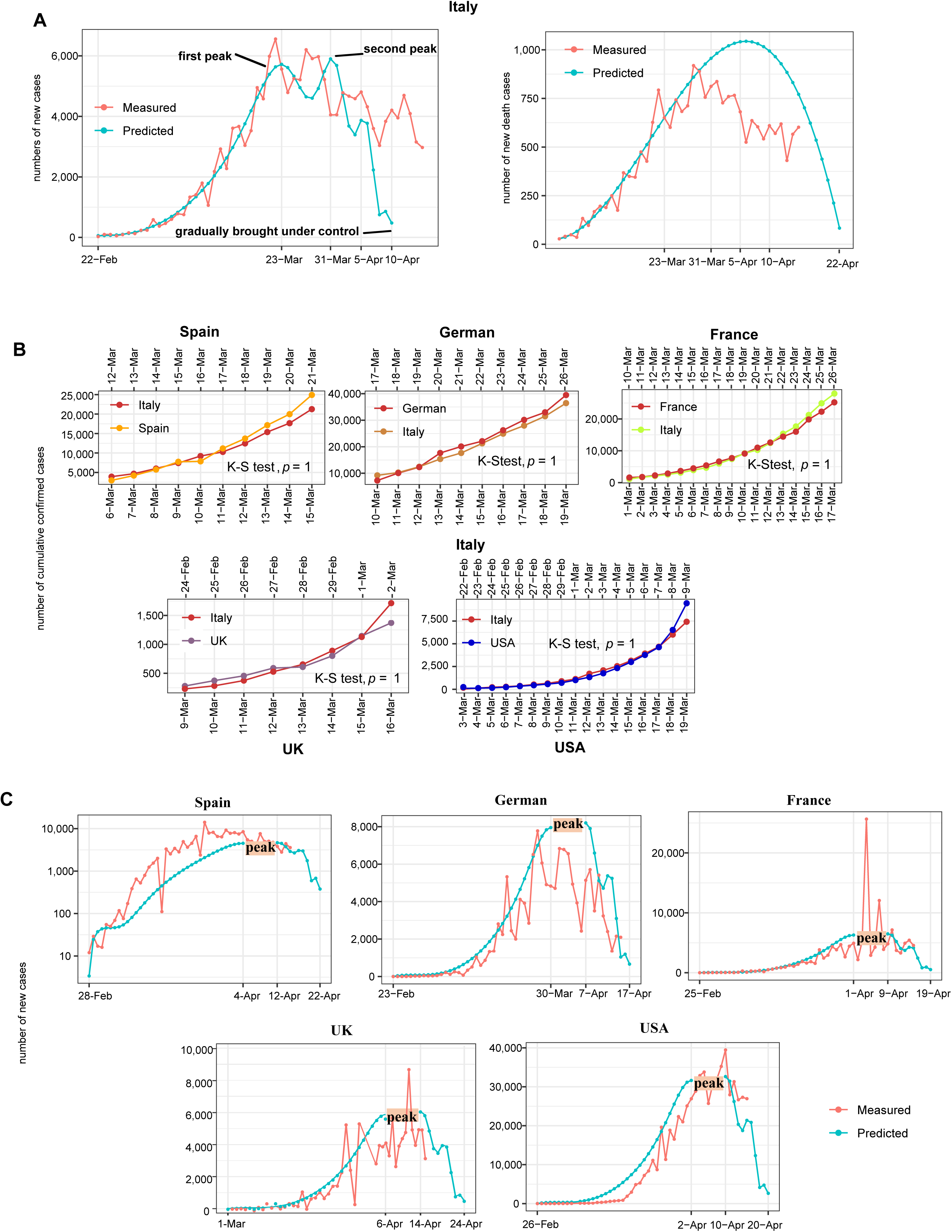
Prediction of the COVID-19 epidemic trend in six other countries. A. Predicted daily numbers of new cases and new death cases of COVID-19 in Italy. B. The numbers of daily new cases in Spain, Germany, France, UK, and USA follow a similar pattern to those in Italy with a 6-, 7-, 9-, 14-, and 10-day lag, respectively (K-S test, *p* = 1). C. Predicted daily numbers of new cases of COVID-19 in Spain, Germany, France, UK, and USA.

Because the COVID-19 epidemics in the other five countries have lagged behind that in Italy and have not shown a turning point, we derived their models from the models for Italy cases. We found that the numbers of daily NCs in Spain, Germany, France, UK, and USA followed a similar pattern to those in Italy with a 6-, 7-, 9-, 14-, and 10-day lag, respectively (K-S test, *p* = 1; Fig. 3B). Accordingly, based on the model to predict the daily number of NCs in Italy, we generated models to predict the daily number of NCs in these countries

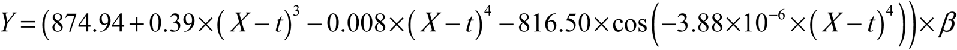

where *Y* is the number of NCs, *X* is the time (days), *t* is the days lagging behind, and *β* is the ratio of the population in that country to that in Italy. For Spain, Germany, France, UK, and USA, *t* was 6, 7, 9, 14, and 10, respectively, and *β* was 0.79, 1.39, 1.10, 1.03, and 5.53, respectively. Based on these models, we predicted that the daily number of NCs would peak between April 4 and April 12, 2020, in Spain, between March 30 and April 7, 2020, in Germany, between April 1 and April 9, 2020, in France, between April 6 and April 14, 2020, in UK, and between April 2 and April 10, 2020, in the USA (Fig. 3C). After around April 20, 2020, the COVID-19 outbreak will be gradually brought under control in these countries. The number of COVID-19 cases is expected to total around 204,000, 360,000, 286, 000, 268,000, and 1,434,000 in Spain, Germany, France, UK, and USA, respectively. Furthermore, we estimated the COVID-19 mortality rate in these countries based on the latest data on April 16, 2020. Likewise, the mortality rate on April 16, 2020, was calculated by dividing the number of DCs on this day by the number of CCCs 10 days before. Accordingly, the COVID-19 mortality rate was 12%, 3.2%, 14.5%, 19.6%, and 6% in Spain, Germany, France, UK, and USA, respectively. Based on these estimated mortality rates, we estimated that around 25,000, 11,500, 41,000, 52,500, and 86,000 people will die of COVID-19 in Spain, Germany, France, UK, and USA, respectively.

## Discussion

The COVID-19 outbreak has made comprehensive and profound impacts on the world. Although this disease appears to be well controlled in China, the recent dramatic increase in new cases and deaths outside China indicates that the outbreak may have catastrophic results globally without implementation of the necessary mitigation measures. However, the experience of China suggests that the outbreak is controllable if effective strategies are employed.^8^ Because the COVID-19 outbreak outside China is in the initial or exponential expansion phase, depending on the region or country, it is currently difficult to predict the major turning points in the COVID-19 epidemic outside China based on their data. Therefore, we assumed that the COVID-19 outbreak outside China follows a similar pattern to that in China, and accordingly derived predictive models for the epidemic outside China from models built based on Chinese cases. Our models predicted that the COVID-19 outbreak would peak across the world around May 22, 2020, and would be basically under control around February 21, 2020 (Fig. 4A). It should be noted that these predictions were made under the premise that countries outside China implement comprehensive and stringent control measures as in China, such as city lockdowns, traffic control, and concentrated medical support for seriously infected areas. Otherwise, the epidemic outside China could follow a different trend, for example, a prolonged outbreak, resulting in more unnecessary deaths and infections. Actually, the COVID-19 outbreak has shown to be more serious than expected in the USA and several Europe countries, including Italy, Spain, France, UK, and Germany, indicating that the early preventive and responsive measures to COVID-19 were insufficient in these countries (Fig. 4B).

**Figure 4.**
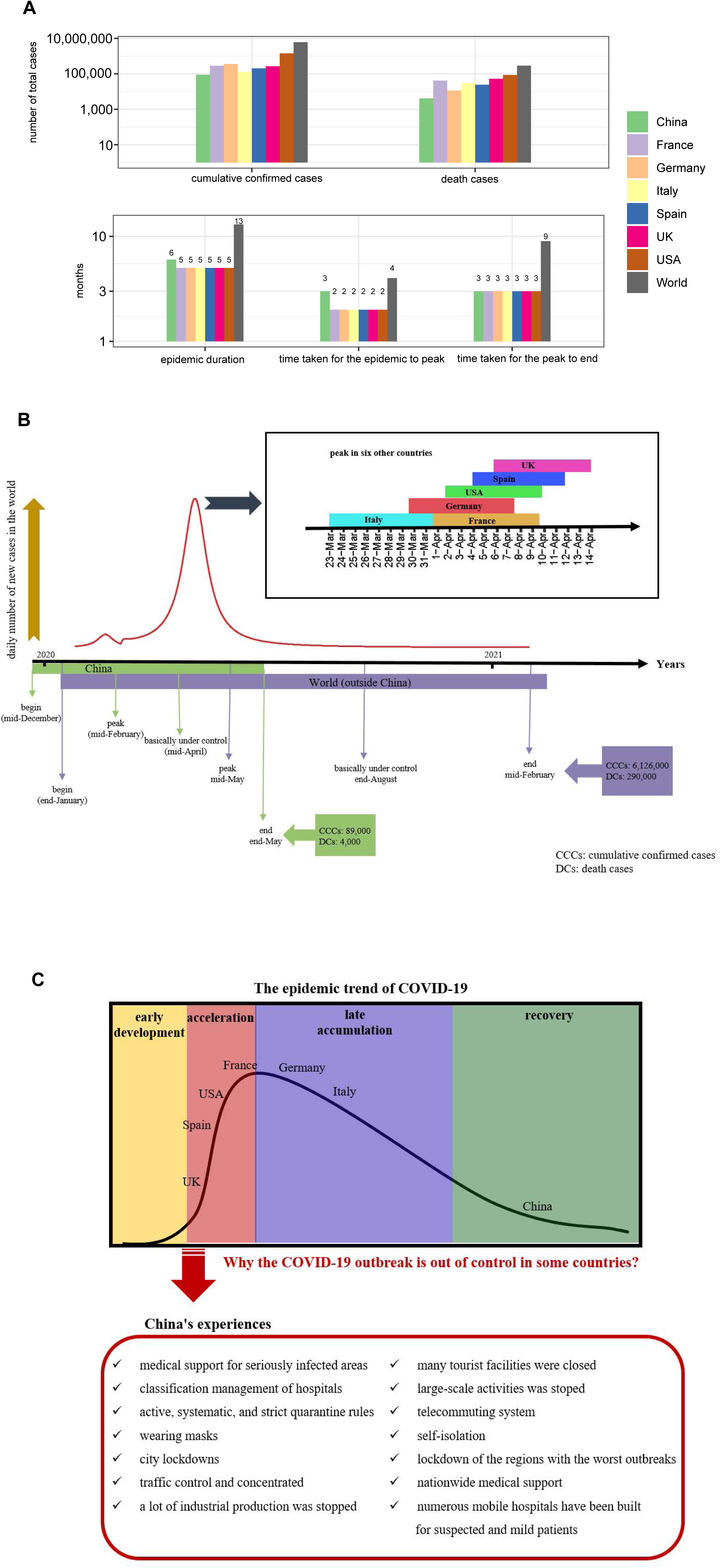
Comparison of the COVID-19 epidemic trend between China, the world outside China, Italy, Spain, Germany, France, UK, and USA. A. The total numbers of cumulative confirmed cases and death cases and epidemic duration. B. The epidemic timelines. C. Comparison of the COVID-19 epidemic trend and the China’s experiences in containing the COVID-19 outbreak.

Our model predicted that the number of NCs would peak on April 9, 2020, outside China. This is consistent with a recent prediction of the epidemic trend of COVID-19 in Italy.^12^ We estimated the mortality rate for COVID-19 to be 4.6% outside China. This number may vary by region and country. For example, the current mortality rate is 12.3% in Italy, compared with 0.1% in Germany. It should be noted that the estimated mortality rate could exceed the actual mortality rate, considering that a number of asymptomatic or mildly-symptomatic cases might not be identified. Actually, a study of 1,099 Chinese cases indicated that the COVID-19 mortality rate in China is 1.4%.^13^ If this is true, then the total number of COVID-19 cases will reach around 230,000, a number far exceeding that reported presently.

The recent rapid spreading of COVID-19 in Europe and the USA highlights the importance of implementing more stringent control measures. Indeed, China’s experiences in containing the spread of COVID-19 can be learnt by other countries currently facing the COVID-19 pandemic (Fig. 4C). For example, China has effectively blocked the massive spreading of COVID-19 by the immediate lockdown of the regions with the worst outbreaks, and reduced more deaths by providing nationwide medical support for these regions. China has implemented the classification management of hospitals with some hospitals concentrating on serving critical COVID-19 cases and others designated for routine medical care. In addition, numerous mobile hospitals have been built to admit suspected and mild patients in China, and strict quarantine rules are enforced. In China, masks are widely used in public area, although the role of face masks in preventing the spread of viruses is debatable.

A limitation of this study is that in deriving predictive models for global cases, we did not take into account factors other than demographics that are associated with the spread and outbreak of COVID-19. These include politics, economy, culture, education, health facilities, geographic position, and race. Overall, the African countries have a low number of COVID-19 cases; the number of cases is also small in the South Asian countries, including India, Pakistan, Bangladesh, and Sri Lanka, despite their high population density. In contrast, the East Asian countries, including China, South Korea, and Japan, have reported a large number of cases, and Europe and USA have replaced China as the centers of the COVID-19 outbreak. The reasons for notably different COVID-19 epidemic sizes in various regions and countries are worth further investigation.

## Conclusions

The COVID-19 outbreak is controllable in the foreseeable future if comprehensive and stringent control measures are taken. Our prediction for world cases assumes that other countries take effective control measures similar to those in China, which would warrant cautious optimism.

## List of abbreviations

CCCs: cumulative confirmed cases
NCs: new cases
DCs: death cases
COVID-19: the corona virus disease 2019
SARS-CoV-2: the 2019 novel coronavirus
K-S test: Kolmogorov-Smirnov test.

## Acknowledgments

We thank LetPub (www.letpub.com) for its linguistic assistance during the preparation of this manuscript.

